# The Challenge Dataset – simple evaluation for safe, transparent healthcare AI deployment

**DOI:** 10.1101/2022.12.15.22280619

**Authors:** James K. Sanayei, Mohamed Abdalla, Monish Ahluwalia, Laleh Seyyed-Kalantari, Simona Minotti, Benjamin A. Fine

## Abstract

In this paper, we demonstrate the use of a “Challenge Dataset”: a small, site-specific, manually curated dataset – enriched with uncommon, risk-exposing, and clinically important edge cases – that can facilitate pre-deployment evaluation and identification of clinically relevant AI performance deficits. The five major steps of the Challenge Dataset process are described in detail, including defining use cases, edge case selection, dataset size determination, dataset compilation, and model evaluation. Evaluating performance of four chest X-ray classifiers (one third-party developer model and three models trained on open-source datasets) on a small, manually curated dataset (410 images), we observe a generalization gap of 20.7% (13.5% - 29.1%) for sensitivity and 10.5% (4.3% - 18.3%) for specificity compared to developer-reported values. Performance decreases further when evaluated against edge cases (critical findings: 43.4% [27.4% - 59.8%]; unusual findings: 45.9% [23.1% - 68.7%]; solitary findings 45.9% [23.1% - 68.7%]). Expert manual audit revealed examples of critical model failure (e.g., missed pneumomediastinum) with potential for patient harm. As a measure of effort, we find that the minimum required number of Challenge Dataset cases is about 1% of the annual total for our site (approximately 400 of 40,000). Overall, we find that the Challenge Dataset process provides a method for local pre-deployment evaluation of medical imaging AI models, allowing imaging providers to identify both deficits in model generalizability and specific points of failure prior to clinical deployment.

## 1. Introduction

Academic institutions and private companies alike are developing AI models designed to interpret medical images, many of which have reported performance that claims to rival or exceed human radiologists.[1,2] However, a growing body of literature suggests that many clinical AI applications fail to generalize in new settings.[3–6] Moreover, healthcare institutions considering deployment of these tools cannot simply assume that the safety of marketed AI models has been verified by regulatory scientists. In some jurisdictions, certain types of predictive software are exempted from such oversight.[7] Additionally, regulatory approval generally implies efficacy - performance under ideal conditions – rather than effectiveness or robustness in the real world. This poses a challenge for intuitions looking to safely adopt AI models, who must operate under a “buyer beware” environment.

## 2. Background

### 2.1. Internal validation

The process of creating an AI model involves splitting a dataset into subsets: one to train the model, and another to validate its performance. However, when models are trained and validated using images from the same source – termed internal validation – the resultant models are vulnerable to biases present in that data that may hamper generalization.[8] One example is sampling bias, such as when medical image datasets from one institution are not representative of the imaging modalities, patient demographics, disease prevalences, and disease state definitions that are present elsewhere.[9,10] Further, bias may be introduced through design choices that may fail to externally generalize, such as site-specific or imprecise labeling of training and validation data,[10] and biases in the local diagnosis and management of diseases.[11] The use of internal validation alone masks these biases, as they are likely to be present in local validation sets but may not reflect the general population. Internal validation therefore often produces unrealistic performance estimates.[8,12,13]

### 2.2. External validation

External validation involves testing a model on data from differing geography, institutions, or practice settings.[8] Numerous literature and lay press examples of classical statistical[4,5] and machine learning models[3,13] show a gap in performance, or “generalization gap”, between internal and external validation. As an extreme example, a deep learning COVID-19 chest X-ray classifier trained on one open-source dataset lost 30 points of AUC when validated on a second dataset.[4] As a result, guidelines such as TRIPOD recommend external validation for predictive model evaluation.[12] Despite this, only 5% of predictive models in the literature report external validation in the abstract or title.[8]

### 2.3. Local validation

While third-party external validation data may shed light on the overall generalizability of a model, it is not necessarily a good predictor of the performance that each healthcare institution will observe upon clinical deployment.[14] In light of the challenges surrounding the deployment of externally-developed models, emerging best practice involves the step of pre-deployment local evaluation.[15,16] Larger academic institutions with sufficient resources may have teams of data scientists with easy access to AI test environments to evaluate a model on local data. However, most healthcare institutions or imaging facilities likely do not have the resources to curate a large local dataset to evaluate performance. Thus, most sites are currently forced to (i) rely on testing data from external scientists or vendors, (ii) forgo local pre-deployment evaluation, or (iii) abandon the idea of adopting AI models.

### 2.4. Edge case testing

In software engineering, edge case testing is a process that involves using extreme, unusual, or otherwise challenging inputs to better validate the behaviour of software. For example, Zhao and Peng (2017)[17] describe an approach for reducing the time and cost involved in validating the reliability of autonomous vehicles by up to 99.9% by “statistically increas[ing] the number of critical driving events” in the evaluation process. In the field of medical imaging, where validating a diagnostic model may require tens of thousands of labeled medical images, edge case testing may provide significant time and cost benefits.

Furthermore, most medical imaging AI models are trained and validated on datasets composed of a fixed number of classes representing observations/diagnoses (e.g., CheXpert[18] contains 14). However, these broad classes frequently contain clinically important subgroups of findings that are not reflected in the labelling schema, leading to hidden performance deficits.[19] They also reflect only a small subset of the full gamut of pathologies encountered in practice, excluding many of the uncommon but important findings present in the “long tail” of imaging findings.[20] Enriching datasets with edge cases may therefore help identify performance deficits that would be missed by most datasets designed through random selection.

### 2.5. Contributions and Significance

This paper i) describes the method for local curation of a site-specific, edge case-enriched external validation dataset (Challenge Dataset) against which outside models can be evaluated to ensure safety (Section: “Materials and Methods”) and ii) demonstrates the use of the Challenge Dataset to evaluate 4 chest X-ray classifiers (Section: “Results”). We also estimate the time required to compile a dataset prospectively to demonstrate its feasibility at most imaging centers.

## 3. Method: Creating a Challenge Dataset

### 3.0. Challenge Dataset Framework

The approach to creating a Challenge Dataset can be outlined in five steps:

1. Determine the intended use of the AI model.
2. Identify a set of applicable edge cases.
3. Determine how many images are needed for each edge case category, given expected model performance and minimum safety requirements.
4. Compile the dataset and assign ground truth labels.
5. Evaluate the proposed model against the dataset by performing both (a) statistical analysis and (b) manual expert audit of discrepant cases.

### 3.1. Step 1: Setting and use cases

The first step in creating a Challenge Dataset is to explicitly decide the intended use of the AI model. This oft-overlooked step includes specifically defining the setting, population, and task being performed (e.g., screening, triage, detection, grading, measurement, diagnosis, prognosis, etc.).[21–23] For radiology, the American College of Radiology – Data Science Institute provides a guide for considering the intended use case.[24]

### 3.2. Step 2: Edge case category selection

Next, to help ensure safe deployment, edge cases should be selected to identify potential gaps in performance overlooked by overall performance measures. Larson et al. (2021)[21] provide an excellent summary of 12 performance elements against which diagnostic models should be evaluated. This includes robust performance against changes in image quality (reliability), modalities and patient populations (applicability), settings (determinism), irrelevant image information (non-distractibility) and elements that promote user trust (fail-safe mechanisms, transparent logic, transparent confidence, ability to be audited/monitored, and an intuitive user interface). In general, it is also useful to include a category consisting of a random selection of images with which to assess the overall generalizability of the model.

There can be no comprehensive nor prescriptive list of edge cases when constructing a Challenge Dataset. While most institutions will have great overlap in their selected edge cases, different use-cases and settings will warrant different edge case testing. For example, the edge cases considered by an oncology center may be much more specialized than those considered by a community practice.

### 3.3. Step 3: Dataset size determination

Having defined the relevant edge cases, the next step is to determine the size of the Challenge Dataset, both overall and for each edge case category. Larger datasets allow for a more precise evaluation of AI models but are more expensive to curate and label. Therefore, determining the minimum required number of images is essential. To help quantify the number of images required, we turn to methods from the external validation of diagnostic models in epidemiology.[25,26]

More specifically, to calculate the number of images required (i.e., the sample size) for our diagnostic test, we use the equations developed found in Appendix 1 of Flahault et al. (2005) and implemented by Matthias Kohl.[27] First, we need to set the *minimal acceptable lower confidence limit –* the lowest performance acceptable to users for a reported Sensitivity (Se) or Specificity (Sp). As defined by Flahault et al. (2005), traditionally the 1−α lower confidence limit for Se (or Sp) can be thought of as the lowest value of Se (or Sp) that is not rejected by a one-sided test of significance level α of the null hypothesis *Se = Se*_*measured*_ (or *Sp = Sp*_*measured*_) against the alternative hypothesis *Se > Se*_*measured*_ (or *Sp > Sp*_*measured*_).

To arrive at the number of images required, using the formulas developed by Flahault et al. (2005), we need: (i) prevalence of the positive class, (ii) expected Se or Sp, (iii) desired power, (iv) significance (taken to be α = 0.05) and (v) the minimal acceptable lower confidence limit. For an implementation, we use the *power.diagnostic.test* function in the MKmisc package which converts these inputs into required sample sizes. In **Table 1** we simulate the required sample size for differing expected performances and minimal acceptable lower confidence limits given the assumptions stated above by varying the expected (i.e., reported sensitivity) and minimal acceptable lower confidence limit but holding all other inputs to the function constant. For an example of how to estimate these variables and determine the required dataset size, please see Step 3 of Results.

**Table 1:** Number of positive cases (or controls) for expected sensitivity (or specificity) ranging from 0.60 to 0.95 to guarantee a minimal acceptable lower confidence limit (ranging from 0.55 to 0.90). Put simply, how many images do you need to have tested to be confident that your model’s performance is below its expected values. For this example, we assume a prevalence of 50%, and choose a power of 0.8 and significance level of 0.05. Created using the R (version 4.1.2) package MKmisc (version 1.8). For an example, refer to Step 3 of Results.

| Reported sensitivity | Minimal acceptable lower confidence limit |  |  |  |  |  |  |  |
| --- | --- | --- | --- | --- | --- | --- | --- | --- |
|  | 0.55 | 0.60 | 0.65 | 0.70 | 0.75 | 0.80 | 0.85 | 0.90 |
| **0.60** | 639 |  |  |  |  |  |  |  |
| **0.65** | 165 | 617 |  |  |  |  |  |  |
| **0.70** | 73 | 157 | 569 |  |  |  |  |  |
| **0.75** | 42 | 71 | 146 | 530 |  |  |  |  |
| **0.80** | 26 | 39 | 62 | 130 | 463 |  |  |  |
| **0.85** | 18 | 24 | 35 | 57 | 117 | 398 |  |  |
| **0.90** | 12 | 17 | 20 | 33 | 50 | 94 | 304 |  |
| **0.95** | 9 | 10 | 12 | 19 | 23 | 37 | 68 | 203 |

### 3.4. Step 4: Dataset compilation

#### 3.4.1 Step 4a: Collect cases

Once the number of images per edge case has been determined, the next step is to compile the Challenge Dataset. A fully manual approach would ask radiologists to prospectively flag cases in their Picture Archive and Communication System (PACS) over a period of time (collected in a database or spreadsheet), which can then be used for AI pre-deployment evaluation. This process can be expedited and facilitated at sites with access to software with report search functionality or a robust “teaching files” case list. Relevant examinations can then be extracted as DICOM images to a Challenge Dataset folder for analysis by local clinicians or analysts in collaboration with the developer/vendor.[28] Since the use of this data is for quality improvement, anonymization may not be required from a research ethics perspective; however, this is best governed by local policies.

#### 3.4.2. Step 4b: Determine ground truth

Finally, ground truth labels must be linked to each Challenge Dataset image. The nature of these labels depends on the possible outputs of the model being evaluated. A binary classifier, for instance, would require binary labels (e.g., “normal” vs “abnormal”).

Various methods can be applied to generate these labels. Ideally, ground truth labels can be added by radiologists at the time that the edge case images are flagged or added to a case list. Otherwise, labels may be retroactively generated by either analyzing the images or the original report text. Review of report text is likely a more economical approach, but this choice introduces a trade-off for error; non-standardized radiology reports often omit findings deemed irrelevant or use inconsistent and imprecise language.[10,29]

### 3.5. Step 5: Model evaluation

The final step is to run the Challenge Dataset images through the AI model and compare model output with the locally collected ground-truth labels.

#### 3.5.1. Step 5a: Quantitative evaluation

Quantitative evaluation involves calculating classification metrics such as sensitivity (recall), specificity, positive predictive value (precision), and negative predictive value. In the case of the “random images” subgroup, used to gauge the overall generalizability of the model, each of these metrics can be calculated and compared to the reported performance. For subgroups that contain only abnormal images (i.e., selected examples of complex, rare, or critical findings), only the sensitivity of the model can be calculated.

It is important to calculate confidence intervals for these values. See Dunnigan, 2008[30] for an example of a very common method for calculating binomial confidence intervals (Clopper-Pearson intervals).

#### 3.5.2. Step 5b: Expert audit

Expert audit refers to a structured audit of examples of model failure by a trained radiologist.[16,31,32] This should involve a detailed review of false positive and false negative cases focusing on identifying i) examples of critical failure that may indicate a patient safety concern, and ii) persistent patterns of failure.

## 4. Results: Application of Framework

### 4.1. Step 1: Setting and use cases

We consider the deployment of a binary chest X-ray model developed by a third party for use by radiologists for triage of chest X-rays in outpatients, inpatients, and emergency department patients. The AI model predicts whether there are any abnormalities in an X-ray image; patients with any abnormality would be triaged for radiologist interpretation before those without. Our study was conducted at Trillium Health Partners, a multi-site community health system in Mississauga, Ontario serving over 1 million patients per year. The candidate model being evaluated was developed by a third party and deployed in an evaluation environment at our institution. The software analyses posteroanterior DICOM chest X-ray images for 10 lung parenchymal, pleural and mediastinal pathologies, and returns a binary output, heat map, and a confidence score. Validation data provided by the third party reports an overall sensitivity of 0.96 and a specificity of 0.93.

### 4.2. Step 2: Edge case category selection

For our specific use case, we concentrate our edge cases on ensuring the robustness and safety of the model. We identified 6 categories of edge cases that are valuable for evaluating specific performance elements outlined by Larson et al. (2021).[21] **Table 2** introduces our categories and explains the rationale for their use. We recommend that local experts identify scenarios in which models may perform unexpectedly, such that their edge cases will either help detect potential performance deficits or provide confidence that model adoption will not introduce risk to patients.

**Table 2:** Composition of each of the six categories of images included in the dataset, as well as the specific AI performance elements they are meant to assess. A total of 250 random images and 160 curated images were acquired. See Appendix A for details.

| Edge Case Category | Number of Images | Properties and Reasoning |
| --- | --- | --- |
| Random images | 250 studies | To test the overall generalizability and applicability to local images. |
| Poor quality | 40 studies (20 normal, 20 abnormal) | Included images noted to be of low quality in radiology reports to test for non-distractibility and reliability. |
| Critical findings | 40 studies (20 random critical findings, 20 select urgent critical findings) | Selected “critical findings” labelled by the radiologist in the report text to test safety and accuracy. |
| Unusual findings | 20 studies | Selected studies containing unusual or uncommon findings from the “long tail” of observations / diagnoses to test the awareness of limitations and fail-safe features of the model as well as safety and accuracy. |
| Solitary findings | 20 studies | Selected studies with only a single finding in them to test for safety and accuracy. |
| Spurious correlates | 40 pneumothorax images (20 with chest tubes and 20 without) | Selected 40 pneumothorax studies to evaluate for non-distractibility (association between chest tube and pneumothorax) |

### 4.3. Step 3: Dataset size determination

To determine the number of images needed per edge case category, we estimated the following variables:

- Prevalence of abnormalities in our patient cohort: this is estimated to be 50% (from prior work at Trillium Health Partners).
- Predicted model performance: this is reported to be approximately 95% both for sensitivity and for specificity (on reported internal validation).
- Minimal acceptable lower confidence limit: for our use case we decided to have multiple minimal acceptable limits for different categories of images. For the general case (i.e., “Random Images”), we selected a value of >0.85 (i.e., able to detect a small drop in performance by having a tight bound). For “Critical Findings” and “Spurious Correlates” (which are likely more difficult than the average case) we chose >0.80 (i.e., a slightly lower confidence limit). For “Unusual Findings” and “Solitary Findings”, we accepted a larger drop in performance; an even looser bound on the lower confidence limit of >0.70 was chosen. For “Poor Quality” images, the lower confidence limit was further reduced to >0.6, as these images could easily be reported by humans as unreliable. **Different scenarios and use cases may require different lower confidence limits**.

Based on these estimates, and using the values in **Table 1**, we would need to collect at least 136 (68*2) images for the “Random Images” category to differentiate between a model performing with the stated performance and one performing below our minimal confidence limit. For “Poor Quality” images we would need 20 images in total (10 positive and 10 negative).

All other categories contain only positively labeled (i.e., abnormal) images and are thus only used to evaluate AI models for sensitivity. For “Critical Findings” and “Spurious Correlates” we would need to curate 40 images per category. For “Unusual Findings” and “Solitary Findings” we would need to curate 19 images per category.

### 4.4. Step 4: Dataset compilation

To compile our dataset, we used Structured Query Language (SQL) to search radiologist reports stored in our local PACS database (Sectra Data Warehouse, Sectra, Sweden). For the Random Images category, binary ground truth labels were generated by manual review of the original report text, and were verified by a board-certified radiologist. Edge case images were also confirmed through a manual review of images by a board-certified radiologist in our PACS. For examples of search terms and details on case-selection strategy, please see Appendix A.

### 4.5. Step 5: Evaluation of third-party developer AI model

#### 4.5.1. Step 5a: Quantitative evaluation

In this section, we use the Challenge Dataset to evaluate the performance of the model and to compare it to the performance reported by the developer. To get a better understanding of model performance relative to existing literature, we also evaluate the performance of 3 chest X-ray classifiers that were trained on open-source datasets on the same set of images as a reference point. More specifically, we evaluate models trained on CheXpert,[18] MIMIC-CXR,[33] and ChestXray14 (termed “NIH” in our results).[34] Full details regarding the implementation of these models is provided in Appendix B.

The results of this analysis can be found in **Figure 1**. 95% confidence intervals were calculated using the Clopper-Pearson method.[30] The first observation is that using a random selection of 250 cases, the developer model does not achieve its reported performance (sensitivity 75.2% [95% CI 66.8% - 82.4%] versus 95.9; specificity 82.9% [95% CI 75.1% - 89.1%] versus 93.4%). The models trained on open-sourced data also demonstrate a generalization gap, showing substantially lower performance than reported.

**Figure 1:**
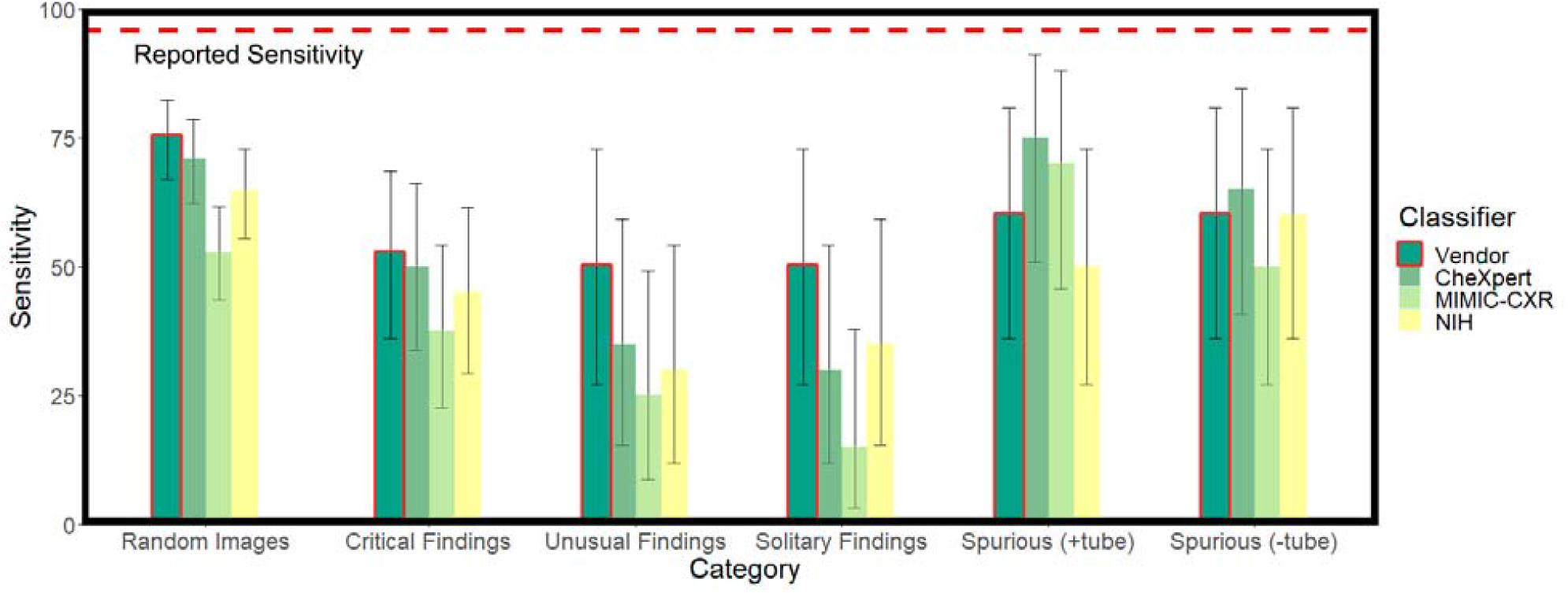
Generalization gap for multiple chest X-ray models. Sensitivity of all four models (one third-party developer and three models trained on open-sourced data for reference) for a subset of the edge cases. The red line indicates the performance of the developer’s model. 95% confidence intervals were calculated using the Clopper-Pearson method.[29] The observed performance of the third-party developer’s model is lower than reported, with the generalization gap decreasing further when tested against edge cases. All results are provided in Appendix D.

Examining the performance of models on our curated edge cases we observe that the generalization gap widens further for all models. Sensitivity for images with critical findings, unusual findings, and solitary findings is only 50-52.5%. This pattern holds across models. The fact that different models, trained on different data sets with different architectures, demonstrate similar performance deficits when applied to our edge cases warrants further investigation.

#### 4.5.2. Step 5b: Expert audit

Our expert audit (performed by 1 radiologist author with 6 years of independent practice experience) compared chest radiographs, radiology reports, and AI predictions for images in the Challenge Dataset. We observed cases with clear implications for clinical deployment. The most striking, presented in **Figure 2**, is that all models failed to detect an obvious case of pneumomediastinum — a finding that can indicate a surgical emergency.

**Figure 2:**
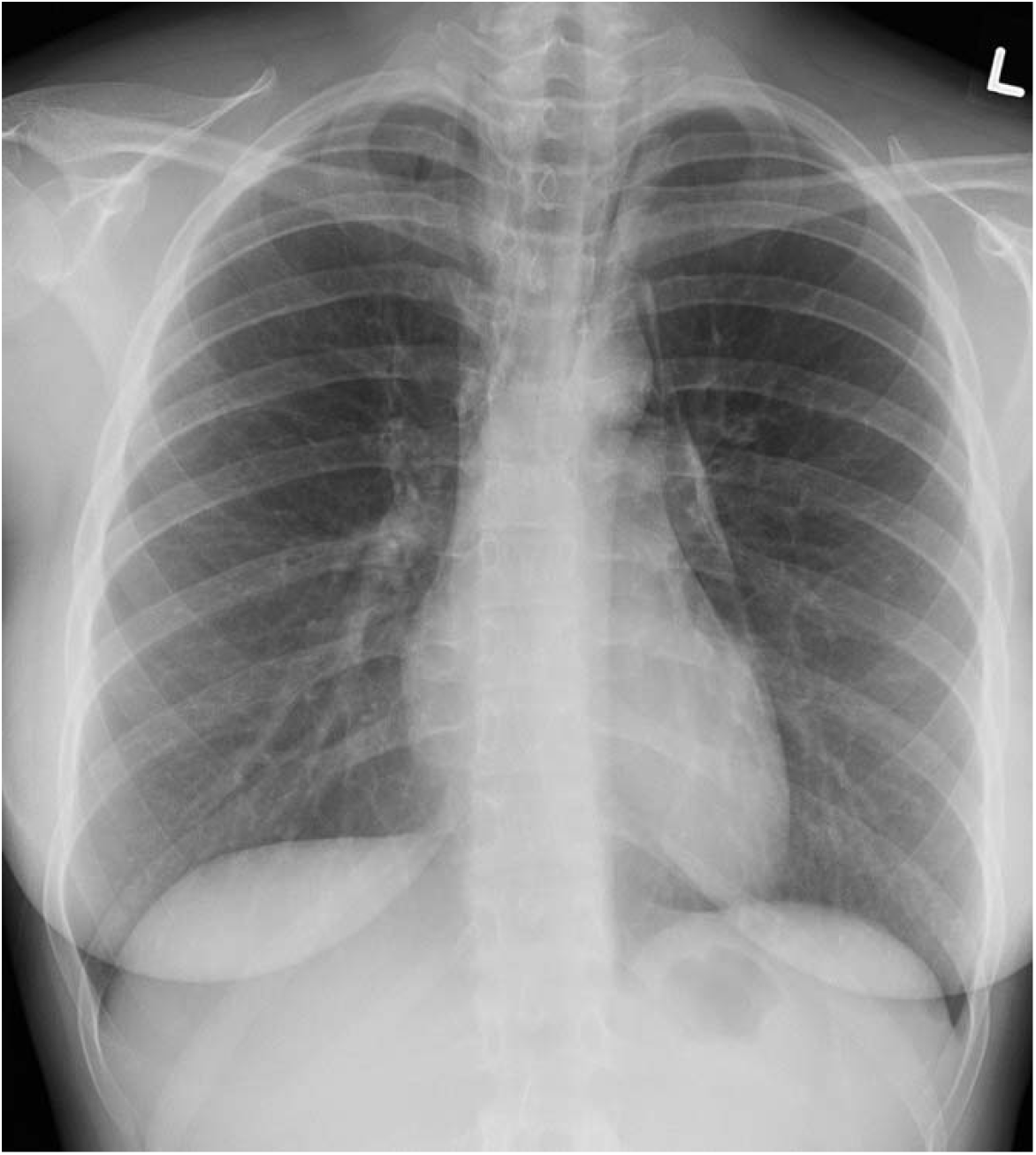
Posteroanterior radiograph in a patient with pneumomediastinum. All models fail to detect this potential surgical emergency. A failure of this magnitude may warrant further edge case testing, retraining, or alteration of deployment plans.

An error of this magnitude might warrant model retraining or redesign. Other examples of missed findings (dextrocardia, bone sclerosis, suboptimal inspiration) may be communicated to users to help them understand the limitations of model performance. In our use case, users would recognize that such cases may or may not be triaged correctly. Finally, some insights may engender confidence and trust in users of the AI model. For instance, the third-party model successfully detected a very small pneumothorax (while 2/3 of other models failed to detect it), and correctly identified a potential new diagnosis of lung cancer (case 6). The full list of results is provided in **Table 3**; the example radiographs with accompanying implications for deployment are provided in Appendix C.

**Table 3:**
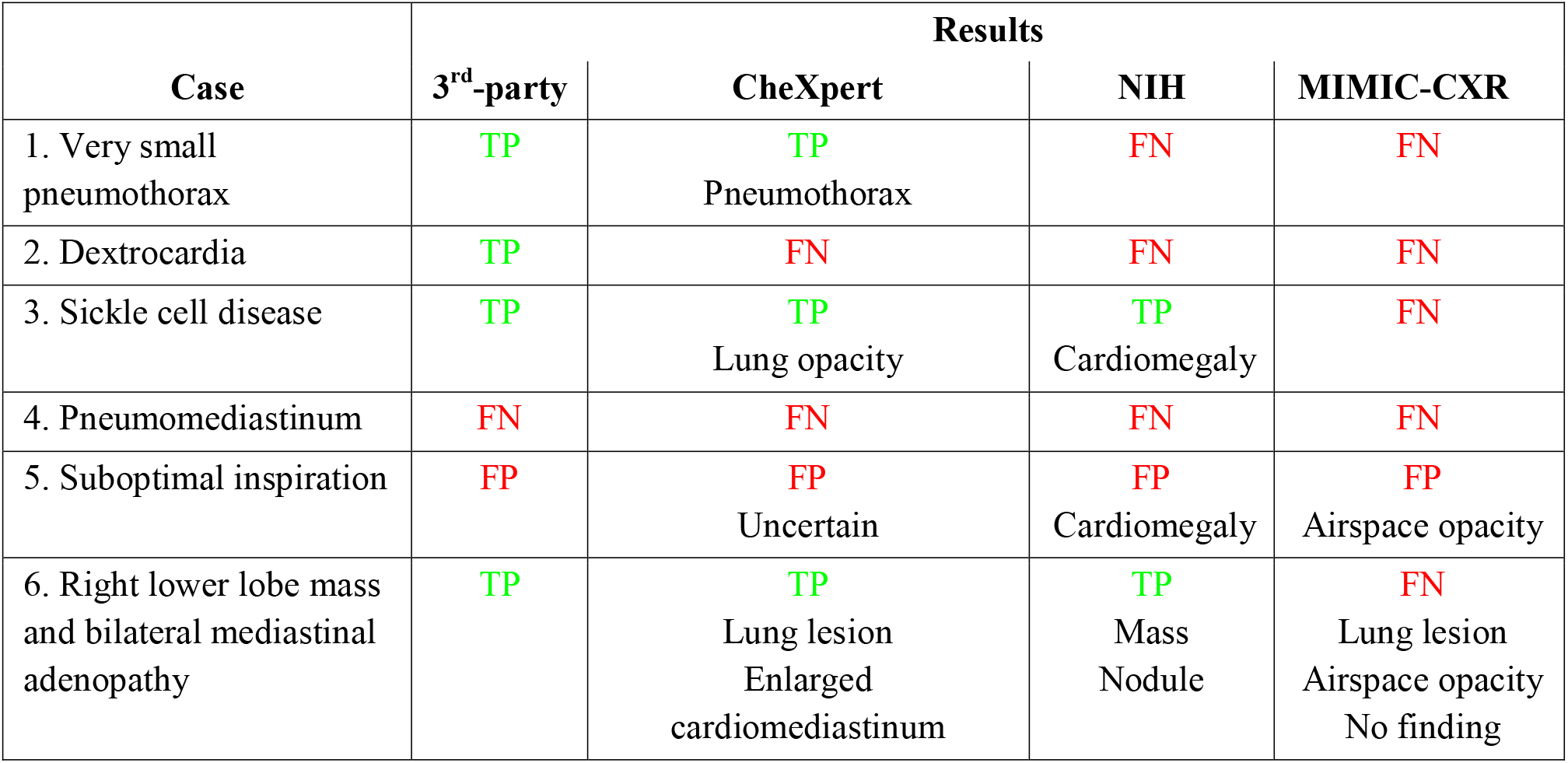
Summary of select example cases from expert audit. For each case, we present the results of the four AI models (TP: true positive, FN: false negative, FP: false positive). Positive results are presented with relevant positive classes for CheXpert, NIH, and MIMIC; the third-party developer software version tested does not provide results by class.

### 4.6 Feasibility

Part of our goal in creating this evaluation process was to enable any site, large or small, to be able to collect requisite images for a local Challenge Dataset. Assuming a site does not routinely archive its medical images and associated reports, it would be necessary to gather images prospectively. As described above, the simplest method is likely for radiologists to flag suitable studies as they are encountered during the delivery of routine care. We assessed the timescale required to create the Challenge Dataset this way (**Figure 3**). In our example, at our institution, over 80% of the 410 images could be collected over a 6-month period, and 96% of this dataset would be created over 1 year. The remaining 4% (from “Poor Quality”, “Critical Findings”, and “Unusual Findings” categories) could be obtained from 3 years of data.

**Figure 3:**
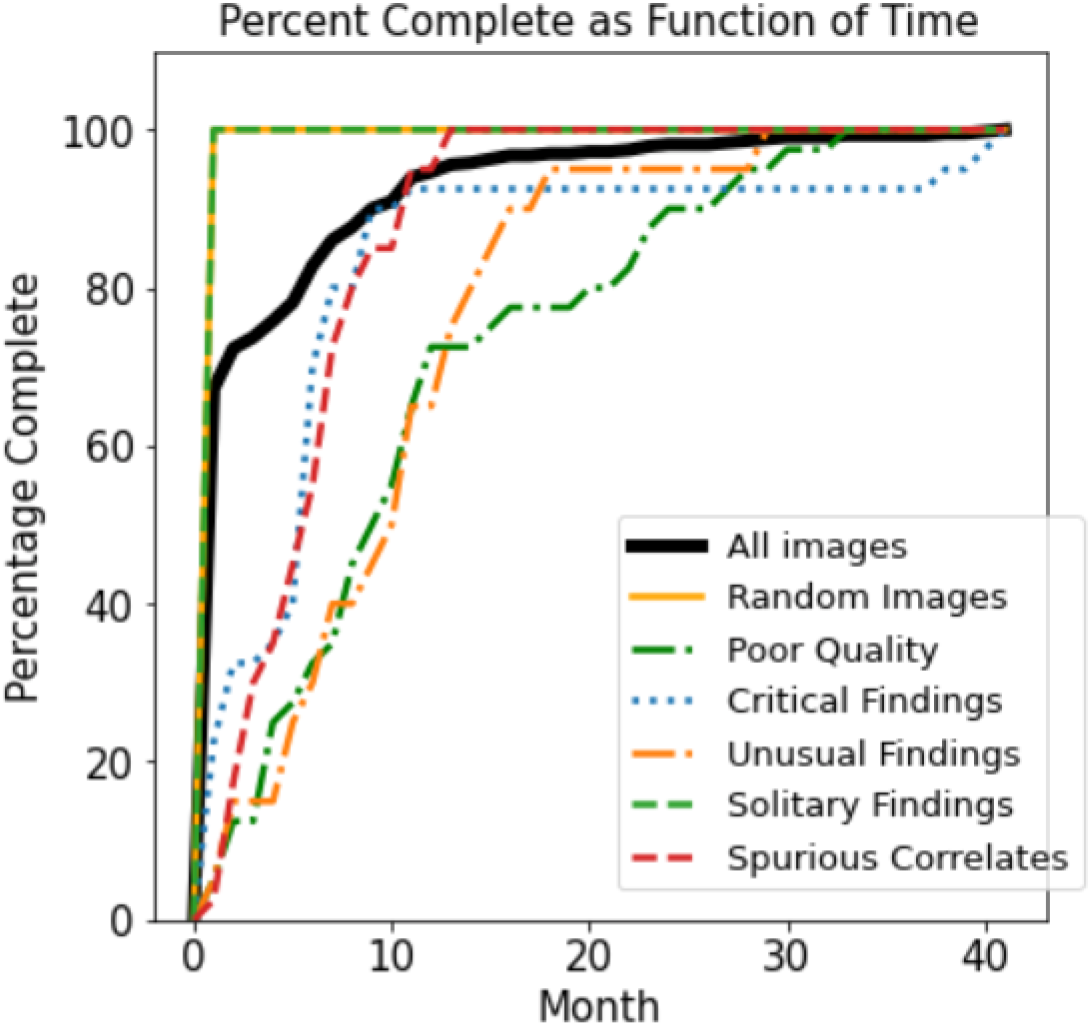
Plot showing the simulated cumulative percentage of a Challenge Dataset collected over time at our institution. This represents the time required to prospectively gather a Challenge Dataset at our institution if radiologists were to collect cases from routine care.

Based on our site volume (approximately 40,000 studies per year) and the size of our dataset (approximately 400 cases), a simple 1%/year rule of thumb emerges, which can be used to estimate the rate at which the dataset can be accumulated. For example, a facility that performs 100,000 studies per year would accumulate 1000 cases for a Challenge Dataset each year (or 400 cases in 5 months). This observation has not yet been validated at other sites.

## 5. Discussion

Pre-deployment evaluation using the Challenge Dataset process described in this work can help bridge the implementation gap in healthcare AI[35] by allowing clinical users to practically and safely manage the deployment of existing AI tools in a number of ways. First and foremost, the Challenge Dataset can be applied easily at any site; imaging providers both large and small can use this approach for cost-effective pre-deployment evaluation to facilitate AI deployment. While more mature sites may employ natural language processing tools to rapidly identify and label a Challenge Dataset at scale, a less well-resourced provider can use a simple spreadsheet to collect data at opportune moments during the course of routine care over a few months.

An additional benefit of curating a Challenge Dataset is that it enables efficiently structured expert audits that can be repeated for many models. Rather than asking experts to explore model performance by randomly sampling images, identifying edge cases a priori helps experts minimize audit time and maximize the information gained from such audits.

Furthermore, insights from Challenge Dataset pre-deployment evaluation can be used by local providers to govern the deployment of AI. For example, poor performance with Spurious Correlates in the setting of chest tubes may lead a provider to decide to “turn off” a model for ICU and inpatients, where the error would be most common. Similarly, gaps in performance identified during pre-deployment Challenge Dataset evaluation can become targets for ongoing monitoring. In our case, for instance, a cross-disciplinary group of clinicians, data scientists, and developers monitoring the deployment of our third-party developer’s model may closely examine its performance in cases with pneumomediastinum, especially as the model is improved. These risks can be summarized and communicated to users in model fact cards.[36]

Another important benefit is that elucidating points of failure can mitigate automation bias. Automation bias — the tendency of humans to rely on automated cues rather than their own judgement — can lead to errors in decision-making.[37,38] For instance, radiologists reading mammography with the assistance of an error-prone computerized detection system were more likely to miss concerning findings than radiologists using no detection system.[39,40] Pre-deployment evaluation with Challenge Datasets can help to reduce the effects of automation bias by providing radiologists with contextual information regarding model failure rates, as well as exposing them to specific examples of model failure (e.g., under detection of solitary findings). Informing users about automation failure behaviour of a system ahead of use has been shown to reduce automation bias.[41,42] More work is required to demonstrate the effect of Challenge Datasets on radiologist performance.

Finally, identifying gaps in performance can raise the level of all future model development and general research. Results from Challenge Dataset evaluations can be published and shared with model developers and other researchers to identify common patterns of failure. These insights can be fed back to improve dataset curation, labeling schema, model development, and software packaging to raise the global quality of healthcare machine learning model development.

## 6. Limitations

There are a number of limitations to this work. First, while we sought to create a process that is replicable, manually gathering and labeling hundreds of medical images may be onerous for some institutions. Software tools to facilitate this process should be considered for future work.

Second, while Challenge Dataset pre-deployment evaluation enables institutions to minimize their image-gathering efforts, smaller datasets result in larger confidence intervals, which may limit the detection of small but significant differences in model performance. For this reason, the Challenge Dataset cannot replace the use of larger datasets for external validation of model performance. It is best used in conjunction with other available data in order to provide a local and manually labeled point of comparison. If potential issues are identified, the smaller size and site-specific nature of the dataset helps to facilitate the review of specific points of failure that can help guide further investigation.

Finally, our selected edge case categories for our proof-of-concept dataset are not comprehensive. For example, we did not seek to stratify performance based on ethnicity, as this data is not routinely collected at our institution, though we would encourage others to do so. Furthermore, for our use-case (triaging), we were primarily concerned with sensitivity, and as such did not gather negative edge cases (apart from negative “Poor Quality” cases) to identify deficits in model specificity.

## 7. Conclusion

Ensuring safe and effective machine learning deployment in healthcare requires rigorous independent pre-deployment external validation. When not performed, models fail to generalize, creating risk for healthcare providers looking to deploy third-party models in their practice. Inspired by edge case testing in automated vehicle safety validation, we show how a small, curated “Challenge Dataset” made up of site-specific and manually labeled images – and artificially enriched with edge cases that can be used to accelerate the evaluation of AI models – can be used to gain clinically meaningful insights into the local performance and critical failure modes of a diagnostic AI model. We also find that creating similar datasets is feasible for most institutions. We hope that the use of the Challenge Dataset approach can enable widespread local validation of medical imaging AI models, enabling safer model deployment and providing feedback to AI developers to globally improve model quality and performance.

## Data Availability

The dataset from this study is held securely at THP. Coded / aggregate data can be made accessible (contact senior author).

## Acknowledgments

We would like to thank Mohannad Hussain for his support with the database and informatics systems. This work would not have been possible without his expertise.

## Author Contributions

BF conceptualized and designed the study. JS acquired the data. All authors assisted in the analysis and interpretation of data. JS drafted the manuscript. BF, MA, MA, LSK, and SM revised the manuscript critically for important intellectual content. All authors gave final approval of the version published.

## Appendix A: Case Selection

To collect images for the “Random Images” category, the most recent 250 posteroanterior chest X-rays in the database were selected. For poor quality, uncertain findings, unusual findings, and spurious correlates, specific search terms were used to isolate images with the desired findings or characteristics. These search terms are outlined in Appendix Table 1 below. For critical findings, the most recent 20 studies performed on Emergency Department patients with the flag “critical finding” in the report were selected. To further enrich the critical findings with less common cases we supplemented this category with an additional 20 images containing select important conditions. In the case of the solitary finding category, images with solitary findings were identified through a manual review of the report text and confirmed by a board-certified radiologist (author). In all categories, more recent images were preferentially selected.

**Appendix Table 1:** Examples of search terms used to compile different edge case categories

| Category | Search terms |
| --- | --- |
| Random images | N/A |
| Poor quality | Quality, suboptimal, rotat%, clothing, motion |
| Critical findings | Critical finding, pneumothorax, pneumomediastinum, metast%, lesion |
| Uncertain findings | Possible, equivocal, borderline, early, subtle |
| Unusual findings | Unusual, herniation, situs inversus, congenital, avascular necrosis, foreign body, bullet, pneumonectomy, mastectomy, aneurysm |
| Solitary findings | N/A |
| Spurious correlates | Pneumothorax, tube |

## Appendix B: Classifier Training

In this work we made use of four different chest X-ray classifiers. The first classifier, and motivation for developing the Challenge Dataset, is a classifier developed by a third party. The third-party developer model was provided as an “out-of-the-box” system; authors did not have access to training or model changes.

To serve as benchmarks, we evaluated three other models trained on open-source datasets: MIMIC-CXR (Johnson et al., 2019), CheXpert (Irvin et al., 2019), and Chest-Xray14 (referred to as NIH) (Wang et al., 2017). A separate classifier was trained on each of the three datasets. The trained classifiers were constructed to perform multi-class classification on each dataset’s available schema. The predictions were then aggregated into a positive and negative class label to mimic the training set-up of the third-party model.

To train the classification models, we made use of the publicly open-sourced code of Seyyed-Kalantari et al. (2020) which can be found online at https://github.com/LalehSeyyed/CheXclusion. We made no modifications to the neural architecture or the hyperparameters of the models described in the associated paper and were able to replicate the same results. We intentionally trained on outside data only to allow for external validation using local data.

## Appendix C: Manual Audit Review

This appendix provides example images from the manual audit of Challenge Dataset cases performed by a board-certified radiologist (author) comparing model predictions to ground truth images/reports. For each example, we present the patient demographics, X-ray image, results from each algorithm and a comment on implications for deployment. These images were selected to help readers understand the powerful and practical insights that can be gleaned from pre-deployment evaluation with a locally curated dataset.

### Case 1

**Appendix Figure 1:**
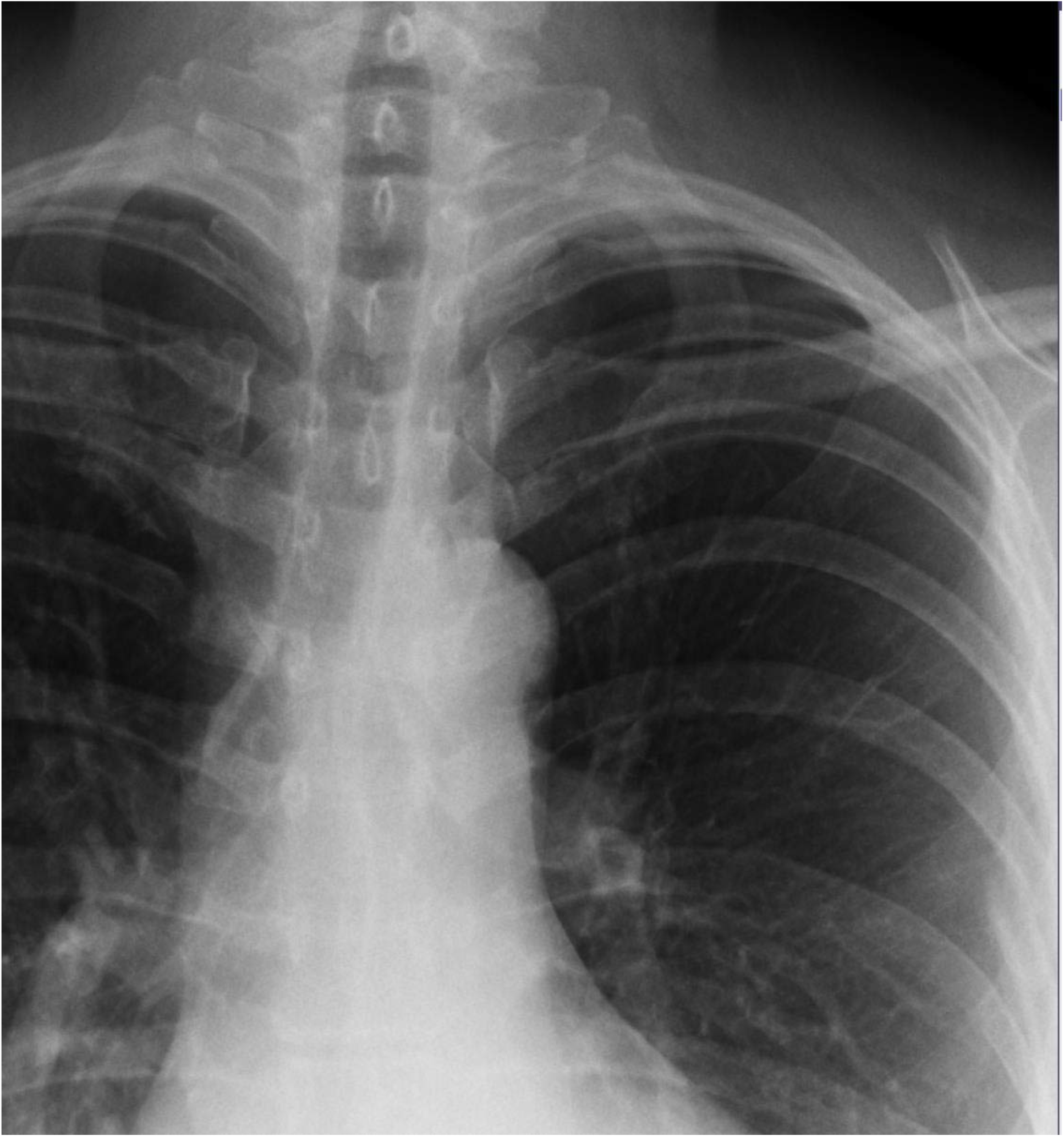
Very small pneumothorax

#### Results

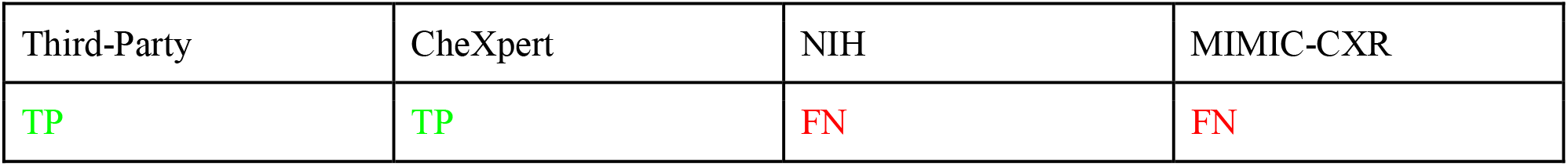

| Third-Party | CheXpert | NIH | MIMIC-CXR |
| --- | --- | --- | --- |
| TP | TP | FN | FN |

#### Implications

Two algorithms (NIH, MIMIC) do not detect the finding. If either of these algorithms were deployed, users would need to know that a normal flag does not exclude a small pneumothorax.

### Case 2

**Appendix Figure 2:**
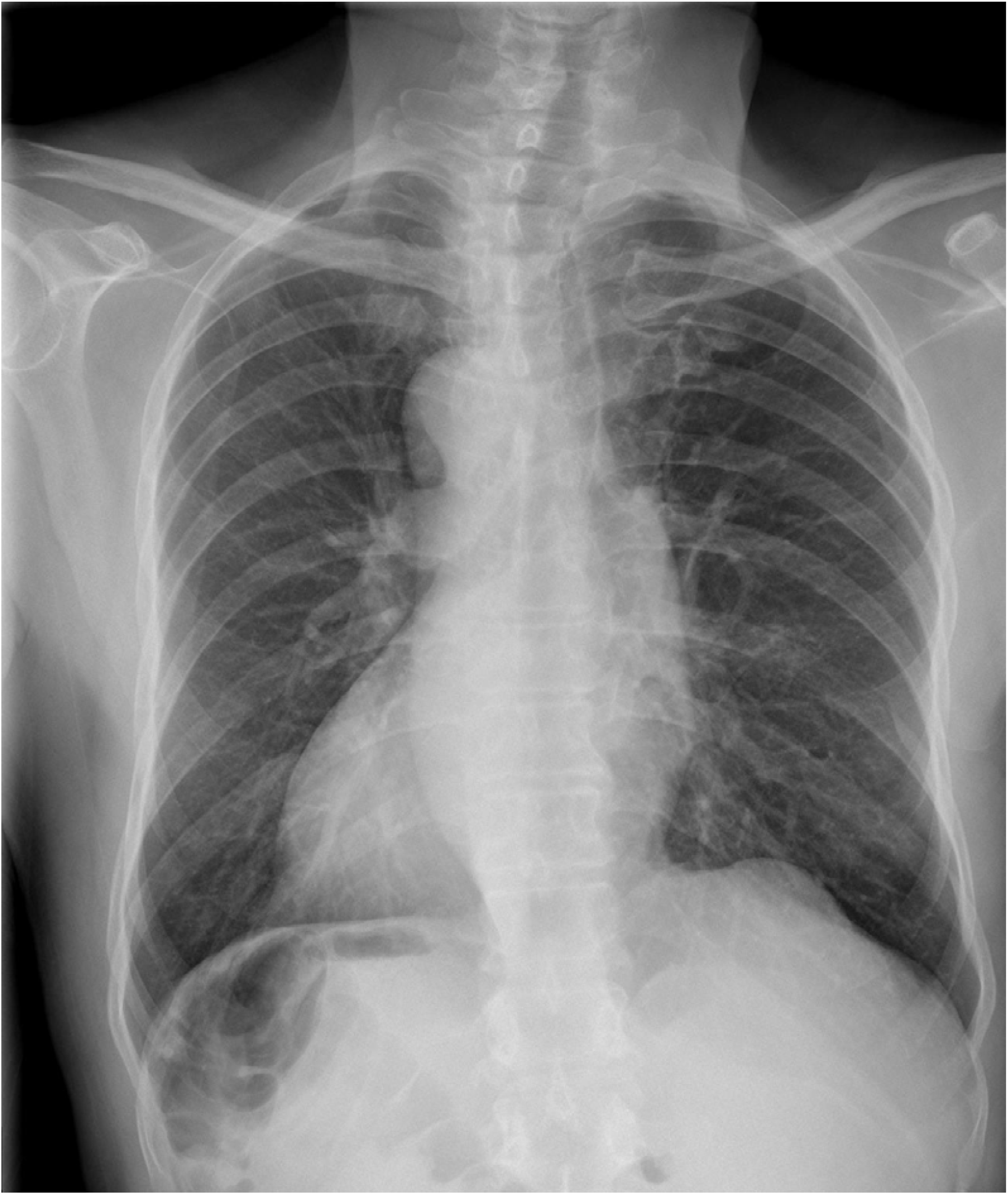
Dextrocardia

#### Results

| Third-Party | CheXpert | NIH | MIMIC-CXR |
| --- | --- | --- | --- |
| TP | FN | FN | FN |

#### Implications

Three algorithms ignore this rare variant and predict a normal examination; one algorithm classifies this as abnormal. This inconsistency points to challenges with labeling schema which are inconsistent and ignore the “long tail” of possible observations. Depending which algorithm was deployed, users would have to be aware that dextrocardia may or may not be characterized as an abnormality.

### Case 3

**Appendix Figure 3:**
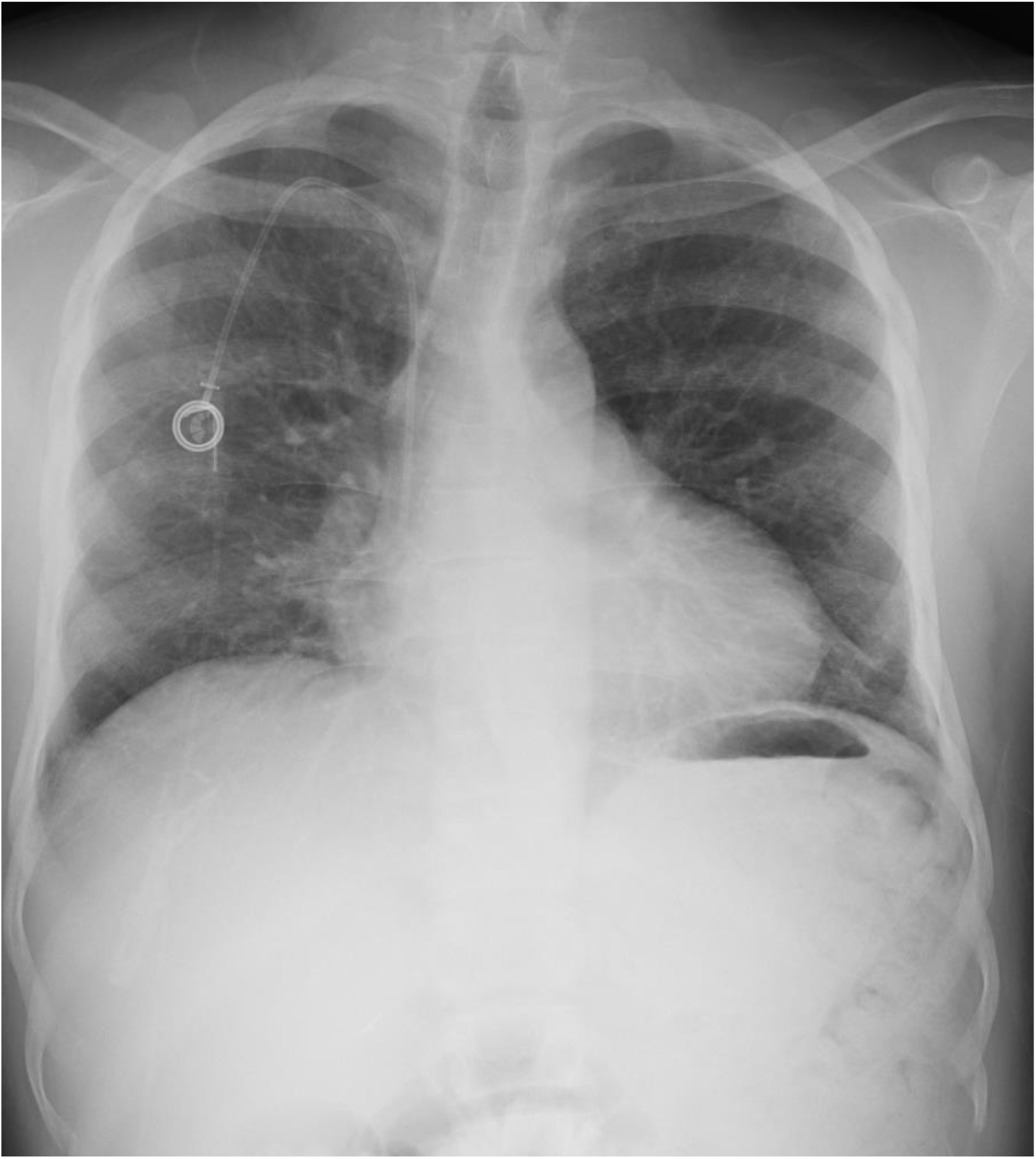
Sickle cell disease

#### Results

| Third-Party | CheXpert | NIH | MIMIC-CXR |
| --- | --- | --- | --- |
| TP | TP<br>“Lung opacity” | TP<br>“Cardiomegaly” | FN |

#### Implications

One algorithm does not detect an abnormality. One detects cardiomegaly. One detects (false positive) lung opacity. None of the algorithms detect the classic finding of diffuse bony sclerosis; bone abnormalities are not within most x-ray algorithm annotation schemas. If deployed, users would need to understand these limitations, which would not be intuitive to most clinical users.

### Case 4

**Appendix Figure 4:**
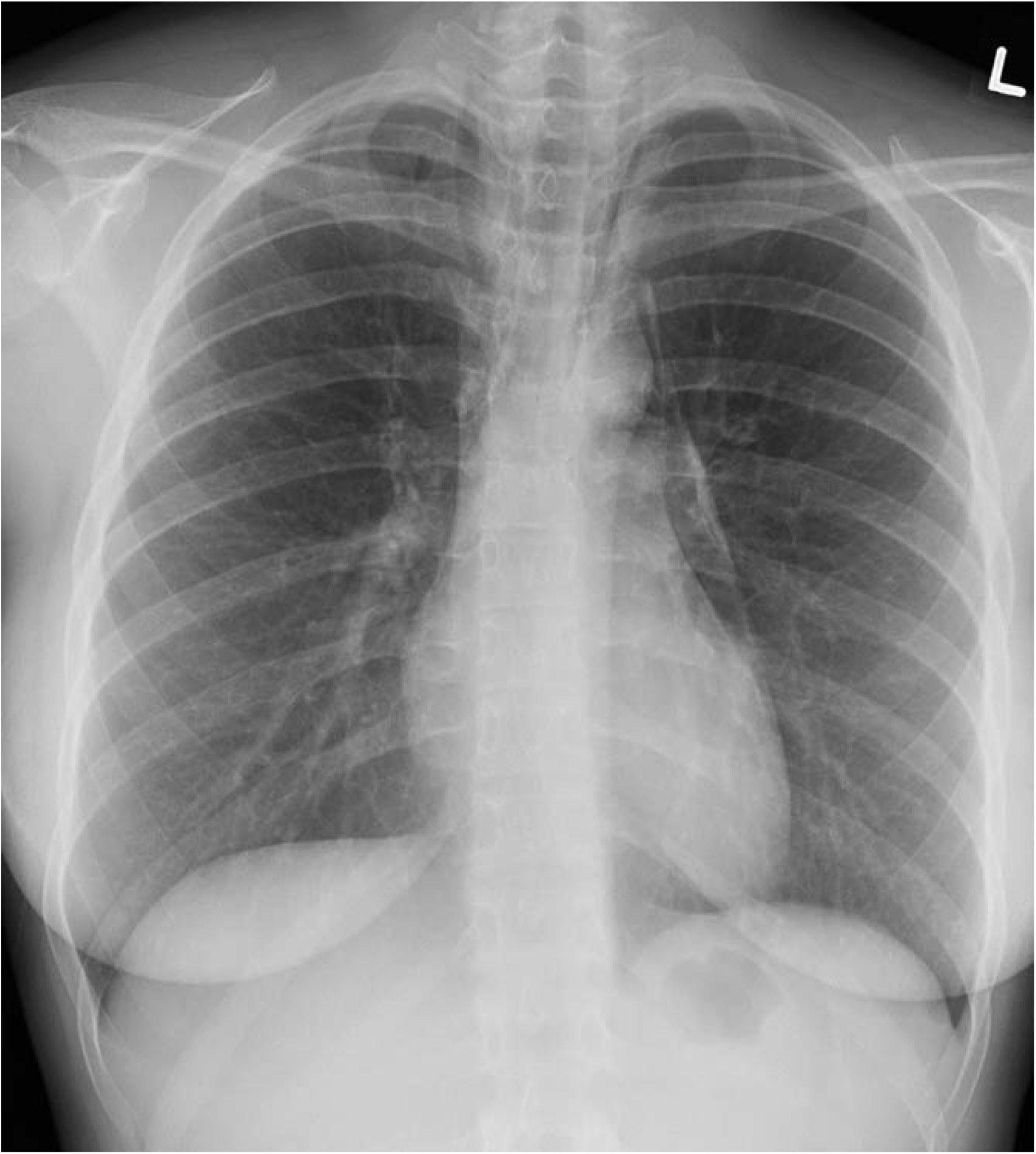
Pneumomediastinum.

#### Results

| Third-Party | CheXpert | NIH | MIMIC-CXR |
| --- | --- | --- | --- |
| FN | FN | FN | FN |

#### Implications

All algorithms fail to detect this potential surgical emergency. A failure of this magnitude may warrant further edge case testing, retraining, or alteration of deployment plans.

### Case 5

**Appendix Figure 5:**
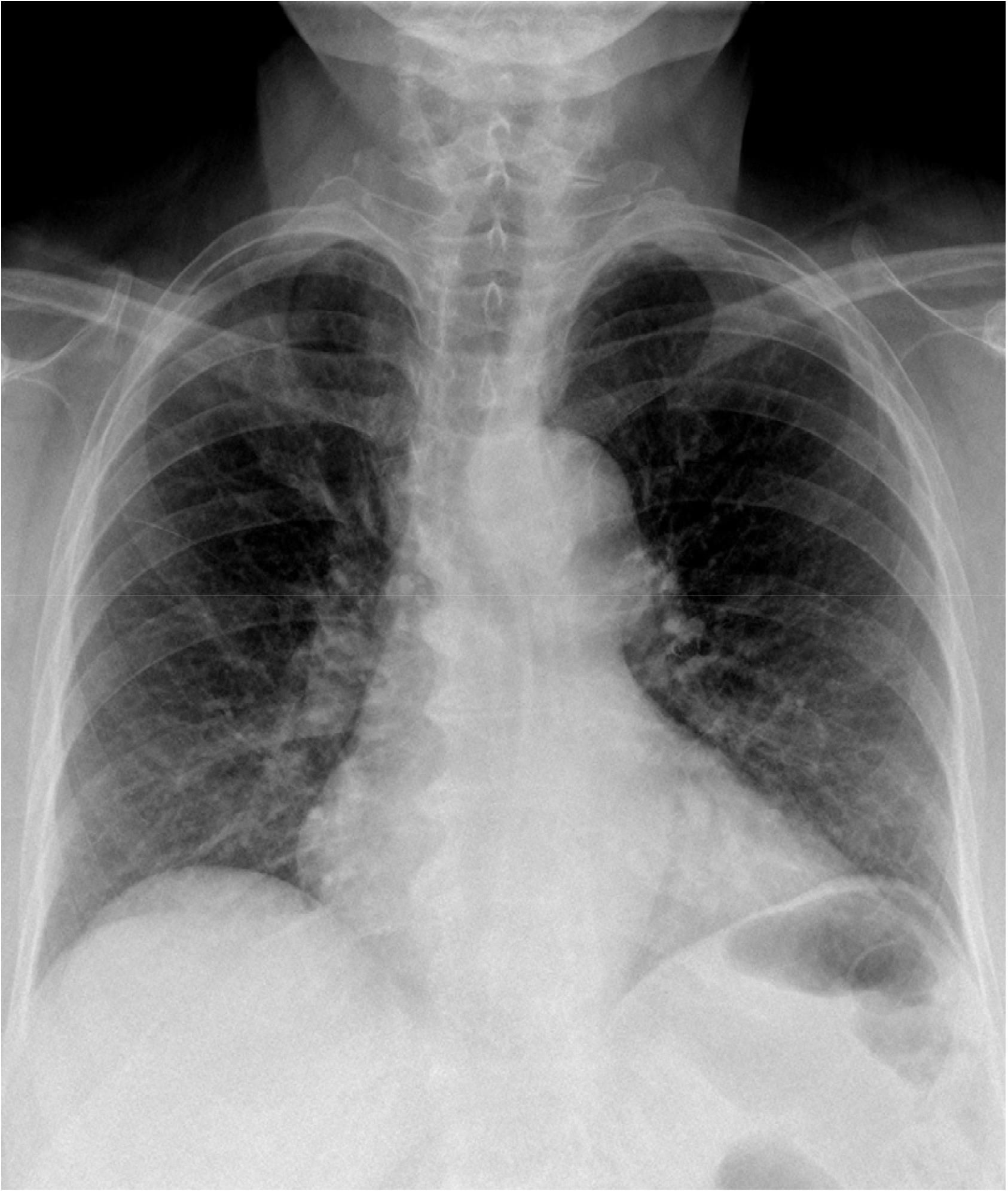
Suboptimal inspiration

#### Results

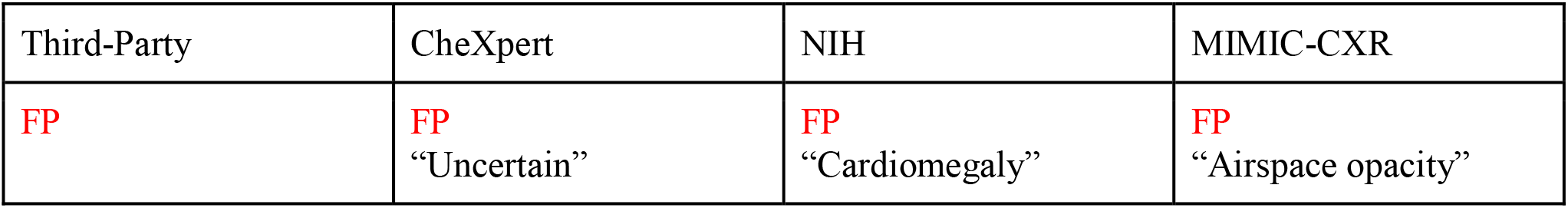

| Third-Party | CheXpert | NIH | MIMIC-CXR |
| --- | --- | --- | --- |
| **FP** | **FP**<br>“Uncertain” | **FP**<br>“Cardiomegaly” | **FP**<br>“Airspace opacity” |

#### Implications

Findings are varied for this “poor quality” case. CheXpert “uncertain” flag is likely the most intuitive result to present to an end user in this scenario.

### Case 6

**Appendix Figure 6:**
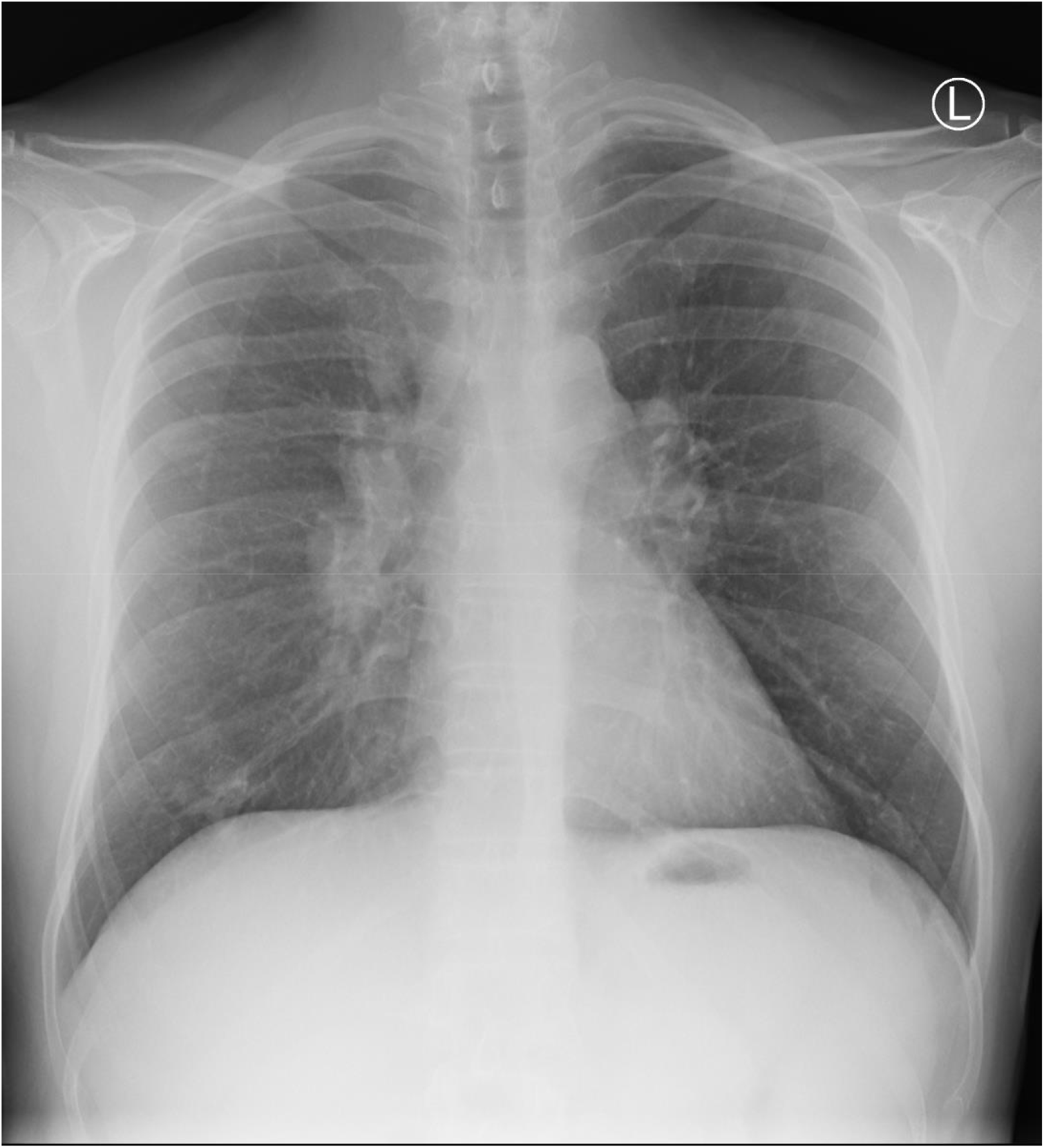
Right lower lobe mass and bilateral mediastinal adenopathy

#### Results

| Third-Party | CheXpert | NIH | MIMIC-CXR |
| --- | --- | --- | --- |
| TP | TP<br>“Lung lesion”<br>“Enlarged<br>cardiomediastinum” | TP<br>“Mass”<br>“Nodule” | FN<br>“Lung lesion”<br>“Airspace opacity”<br>“No finding” |

#### Implications

All algorithms detect the lung abnormality, building user trust. Only 1 model detects the marked mediastinal lymphadenopathy. Curiously, MIMIC also triggers the “no finding” class, which leads to a FN interpretation in our deployment – a potential source of confusion for users and deployment team.

## Appendix D: Model Performance Tables

**Appendix Table 2:** Performance of the **third-party developer** provided model when evaluated on the curated Challenge Dataset. Reported sensitivity: 95.9. Reported specificity: 93.4. 95% confidence intervals provided in brackets determined by the Clopper-Pearson method (Dunnigan, 2008)

| Category | Sensitivity / Recall | Specificity | PPV / Precision | NPV |
| --- | --- | --- | --- | --- |
| Random Images | 75.2<br>(66.8 - 82.4) | 82.9<br>(75.1 - 89.1) | 81.4<br>(74.6 - 86.8) | 76.7<br>(70.7 - 81.8) |
| Poor Quality | 80.0<br>(56.3 - 94.3) | 75.0<br>(50.9 - 91.3) | 76.2<br>(59.2 - 87.6) | 79.0<br>(60.1 - 90.3) |
| Critical Findings | 52.5<br>(36.1 - 68.5) | - | - | - |
| Unusual Findings | 50<br>(27.2 - 72.8) | - | - | - |
| Solitary Findings | 50<br>(27.2 - 72.8) | - | - | - |
| Spurious Correlates<br>(with tube) | 60.0<br>(36.0 - 80.9) | - | - | - |
| Spurious Correlates<br>(without tube) | 60.0<br>(36.0 - 80.9) | - | - | - |

**Appendix Table 3:** Performance of the **CheXpert** model when evaluated on the curated Challenge Dataset. 95% confidence intervals provided in brackets determined by the Clopper-Pearson method (Dunnigan, 2008)

| Category | Sensitivity / Recall | Specificity | PPV / Precision | NPV |
| --- | --- | --- | --- | --- |
| Random Images | 70.9<br>(62.2 - 78.6) | 87.0<br>(79.7 - 92.4) | 84.5<br>(77.3 - 89.7) | 74.9<br>(69.3 - 79.8) |
| Poor Quality | 70.0<br>(45.7 - 88.1) | 65.0<br>(40.8 - 84.6) | 66.7<br>(50.8 - 79.5) | 68.4<br>(50.8 - 82.0) |
| Critical Findings | 50.0<br>(33.8 - 66.2) | - | - | - |
| Unusual Findings | 35.0<br>(15.4 - 59.2) | - | - | - |
| Solitary Findings | 30.0<br>(11.9 - 54.3) | - | - | - |
| Spurious Correlates<br>(with tube) | 75.0<br>(50.9 - 91.3) | - | - | - |
| Spurious Correlates<br>(without tube) | 65.0<br>(40.8 - 84.6) | - | - | - |

**Appendix Table 4:**
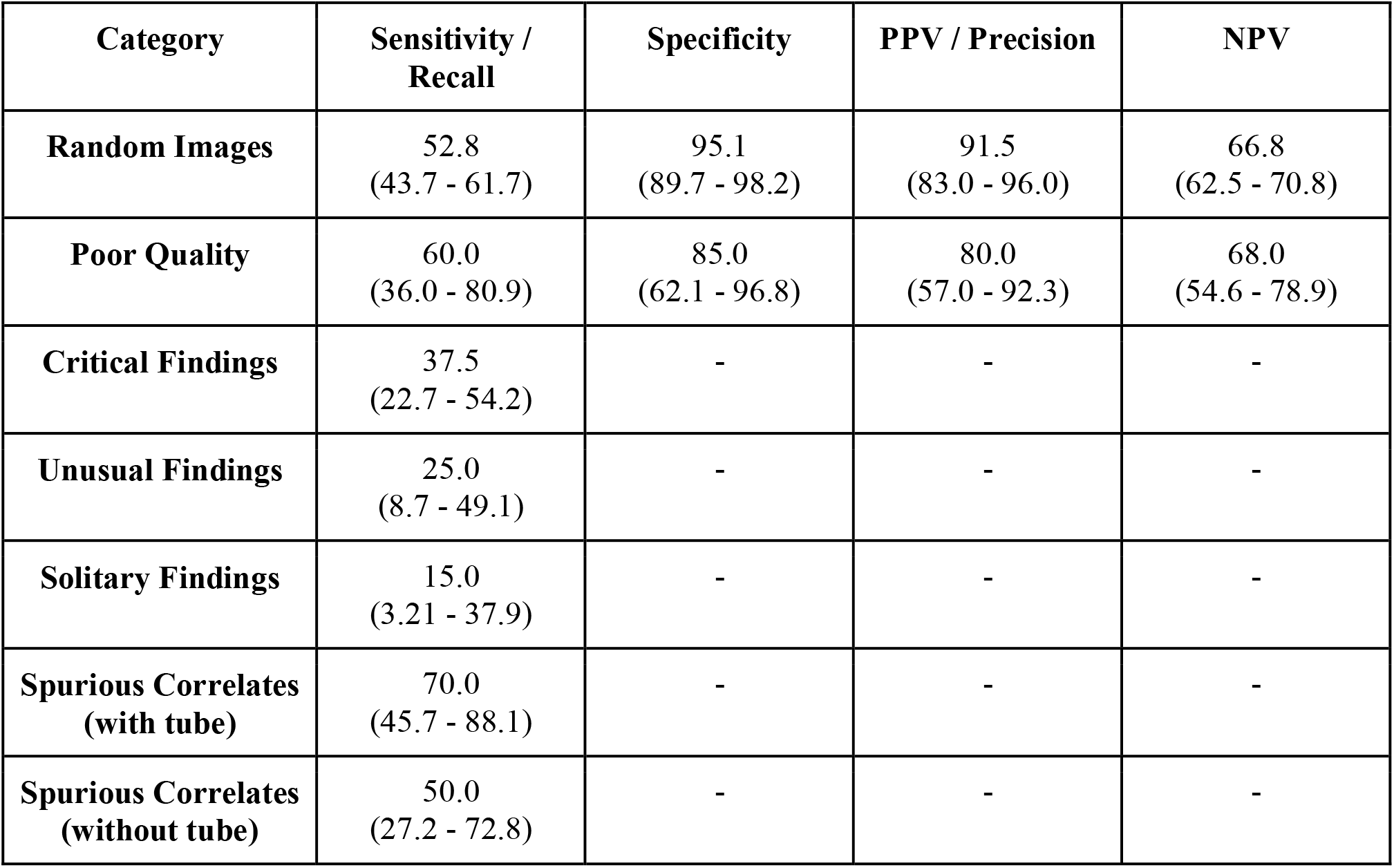
Performance of the **MIMIC-CXR** model when evaluated on the curated Challenge Dataset. 95% confidence intervals provided in brackets determined by the Clopper-Pearson method (Dunnigan, 2008)

**Appendix Table 5:**
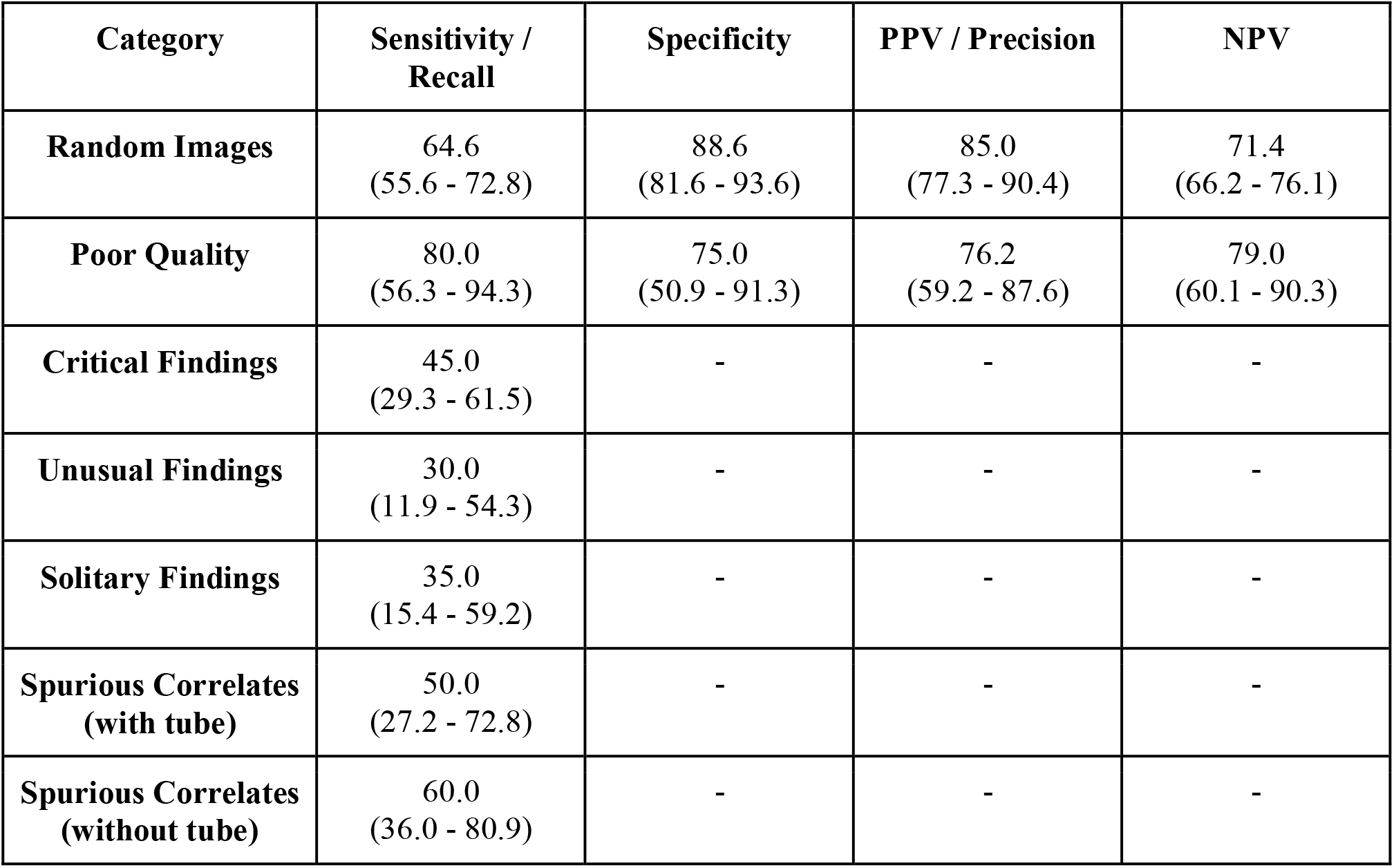
Performance of the **NIH** model when evaluated on the curated Challenge Dataset. 95% confidence intervals provided in brackets determined by the Clopper-Pearson method (Dunnigan, 2008)

| Category | Sensitivity / Recall | Specificity | PPV / Precision | NPV |
| --- | --- | --- | --- | --- |
| Random Images | 64.6<br>(55.6 - 72.8) | 88.6<br>(81.6 - 93.6) | 85.0<br>(77.3 - 90.4) | 71.4<br>(66.2 - 76.1) |
| Poor Quality | 80.0<br>(56.3 - 94.3) | 75.0<br>(50.9 - 91.3) | 76.2<br>(59.2 - 87.6) | 79.0<br>(60.1 - 90.3) |
| Critical Findings | 45.0<br>(29.3 - 61.5) | - | - | - |
| Unusual Findings | 30.0<br>(11.9 - 54.3) | - | - | - |
| Solitary Findings | 35.0<br>(15.4 - 59.2) | - | - | - |
| Spurious Correlates<br>(with tube) | 50.0<br>(27.2 - 72.8) | - | - | - |
| Spurious Correlates<br>(without tube) | 60.0<br>(36.0 - 80.9) | - | - | - |

## References

1 Rajpurkar P, Irvin J, Zhu K, et al. CheXNet: Radiologist-Level Pneumonia Detection on Chest X-Rays with Deep Learning. arXiv Prepr Published Online First: 2017.

2 Wu JT, Wong KCL, Gur Y, et al. Comparison of Chest Radiograph Interpretations by Artificial Intelligence Algorithm vs Radiology Residents. JAMA Netw Open 2020;3:e2022779–e2022779.

3 Wong A, Otles E, Donnelly JP, et al. External validation of a widely implemented proprietary sepsis prediction model in hospitalized patients. JAMA Intern Med 2021;181:1065–70.

4 DeGrave AJ, Janizek JD, Lee SI. AI for radiographic COVID-19 detection selects shortcuts over signal. Nat Mach Intell 2021;3:610–9.

5 Siontis GCM, Tzoulaki I, Castaldi PJ, et al. External validation of new risk prediction models is infrequent and reveals worse prognostic discrimination. J Clin Epidemiol 2015;68:25–34.

6 Zhang H, Dullerud N, Seyyed-Kalantari L, et al. An empirical framework for domain generalization in clinical settings. In: ACM CHIL 2021 - Proceedings of the 2021 ACM Conference on Health, Inference, and Learning. Association for Computing Machinery, Inc 2021. 279–90.

7 US Food and Drug Administration (FDA). FDA Changes to existing medical software policies resulting from section 3060 of the 21st century cures Act. 2019.

8 Ramspek CL, Jager KJ, Dekker FW, et al. External validation of prognostic models: what, why, how, when and where? Clin Kidney J 2021;14:49–58.

9 US Food and Drug Administration (FDA). Artificial Intelligence/Machine Learning (AI/ML)-Based Software as a Medical Device (SaMD) Action Plan. 2021.

10 Willemink MJ, Koszek WA, Hardell C, et al. Preparing medical imaging data for machine learning. Radiology 2020;295:4–15.

11 Geis JR, Brady AP, Wu CC, et al. Ethics of artificial intelligence in radiology: Summary of the joint European and North American multisociety statement. Radiology 2019;293:436–40.

12 Moons KGM, Altman DG, Reitsma JB, et al. Transparent reporting of a multivariable prediction model for individual prognosis or diagnosis (TRIPOD): Explanation and elaboration. Ann Intern Med 2015;162:W1–73.

13 Yu AC, Mohajer B, Eng J. External Validation of Deep Learning Algorithms for Radiologic Diagnosis: A Systematic Review. Radiol Artif Intell 2022;4.

14 Futoma J, Simons M, Panch T, et al. The myth of generalisability in clinical research and machine learning in health care. Lancet Digit Heal 2020;2:e489–92.

15 Allen B, Dreyer K, Stibolt R, et al. Evaluation and Real-World Performance Monitoring of Artificial Intelligence Models in Clinical Practice: Try It, Buy It, Check It. J Am Coll Radiol 2021;18:1489–96.

16 Mahajan V, Venugopal VK, Murugavel M, et al. The Algorithmic Audit: Working with Vendors to Validate Radiology-AI Algorithms—How We Do It. Acad Radiol 2020;27:132–5.

17 Zhao D, Peng H. From the Lab to the Street: Solving the Challenge of Accelerating Automated Vehicle Testing. arXiv Prepr Published Online First: 15 July 2017.

18 Irvin J, Rajpurkar P, Ko M, et al. CheXpert: A large chest radiograph dataset with uncertainty labels and expert comparison. In: 33rd AAAI Conference on Artificial Intelligence, AAAI 2019, 31st Innovative Applications of Artificial Intelligence Conference, IAAI 2019 and the 9th AAAI Symposium on Educational Advances in Artificial Intelligence, EAAI 2019. 2019. 590–7.

19 Oakden-Rayner L, Dunnmon J, Carneiro G, et al. Hidden stratification causes clinically meaningful failures in machine learning for medical imaging. In: ACM CHIL 2020 - Proceedings of the 2020 ACM Conference on Health, Inference, and Learning. 2020. 151–9.

20 Kahn CE. The Long Tail. Radiol Artif Intell 2019.

21 Larson DB, Harvey H, Rubin DL, et al. Regulatory Frameworks for Development and Evaluation of Artificial Intelligence–Based Diagnostic Imaging Algorithms: Summary and Recommendations. J Am Coll Radiol 2021;18:413–24.

22 Omoumi P, Ducarouge A, Tournier A, et al. To buy or not to buy—evaluating commercial AI solutions in radiology (the ECLAIR guidelines). Eur Radiol 2021;31:3786–96.

23 Tang A, Tam R, Cadrin-Chênevert A, et al. Canadian Association of Radiologists White Paper on Artificial Intelligence in Radiology. Can Assoc Radiol J 2018;69:120–35.

24 American College of Radiology Data Science Institute. Define-AI Directory.

25 Bujang MA, Adnan TH. Requirements for minimum sample size for sensitivity and specificity analysis. J Clin Diagnostic Res 2016;10:YE01–6.

26 Flahault A, Cadilhac M, Thomas G. Sample size calculation should be performed for design accuracy in diagnostic test studies. J Clin Epidemiol 2005;58:859–62.

27 Kohl M. MKmisc: Miscellaneous functions from M. Kohl. R package version 0.91. 2012.

28 Lui YW, Geras K, Block KT, et al. How to Implement AI in the Clinical Enterprise: Opportunities and Lessons Learned. J Am Coll Radiol 2020;17:1394–7.

29 Pons E, Braun LMM, Hunink MGM, et al. Natural language processing in radiology: A systematic review. Radiology 2016;279:329–43.

30 Dunnigan K. Confidence interval for Binomial Proportions. In: MWSUG Conference, Indianapolis, IN. 2008.

31 Garbin C, Marques O. Assessing Methods and Tools to Improve Reporting, Increase Transparency, and Reduce Failures in Machine Learning Applications in Health Care. Radiol Artif Intell 2022;4.

32 Liu X, Glocker B, McCradden MM, et al. The medical algorithmic audit. Lancet Digit Heal 2022;4:e384–97.

33 Johnson AEW, Pollard TJ, Berkowitz SJ, et al. MIMIC-CXR, a de-identified publicly available database of chest radiographs with free-text reports. Sci Data 2019;6.

34 Wang X, Peng Y, Lu L, et al. ChestX-ray8: Hospital-scale chest X-ray database and benchmarks on weakly-supervised classification and localization of common thorax diseases. In: Proceedings - 30th IEEE Conference on Computer Vision and Pattern Recognition, CVPR 2017. Institute of Electrical and Electronics Engineers Inc. 2017. 3462–71.

35 Seneviratne MG, Shah NH, Chu L. Bridging the implementation gap of machine learning in healthcare. BMJ Innov 2020;6:45–7.

36 Sendak MP, Gao M, Brajer N, et al. Presenting machine learning model information to clinical end users with model facts labels. npj Digit Med 2020;3:1–4.

37 Magrabi F, Ammenwerth E, McNair JB, et al. Artificial Intelligence in Clinical Decision Support: Challenges for Evaluating AI and Practical Implications. Yearb Med Inform 2019;28:128–34.

38 Lyell D, Coiera E. Automation bias and verification complexity: a systematic review. J. Am. Med. Inform. Assoc. 2017;24:423.

39 Alberdi E, Povyakalo A, Strigini L, et al. Effects of incorrect computer-aided detection (CAD) output on human decision-making in mammography. Acad Radiol 2004;11:909–18.

40 Povyakalo AA, Alberdi E, Strigini L, et al. How to discriminate between computer-aided and computer-hindered decisions: A case study in mammography. Med Decis Mak 2013;33:98–107.

41 Bagheri N, Jamieson GA. The impact of context-related reliability on automation failure detection and scanning behaviour. In: Conference Proceedings - IEEE International Conference on Systems, Man and Cybernetics. 2004. 212–7.

42 Bahner JE, Hüper AD, Manzey D. Misuse of automated decision aids: Complacency, automation bias and the impact of training experience. Int J Hum Comput Stud 2008;66:688–99.

